# Levels and trends in fertility rates among adolescents aged 15–19 in 21 States and Union Territories in India from 1990 to 2050: A Bayesian Modelling Study

**DOI:** 10.64898/2025.12.17.25342447

**Authors:** Fengqing Chao, Yifei Su, Christophe Z. Guilmoto

## Abstract

**Objectives:** Adolescent fertility in India remains underexplored at the state level. This study estimates age-specific fertility rates (ASFRs) for females aged 15–19 across all major Indian states and union territories (UTs) from 1990 to 2025 and generates probabilistic projections of state-level ASFRs for females aged 15–19 through 2050.

**Design:** Bayesian modelling based on fertility data from five phases of the India National Family Health Surveys (NFHSs) in 1992–93, 1998–99, 2005–06, 2015–16, and 2019–21 and the Sample Registration System from 1971 to 2007.

**Setting:** India.

**Participants:** (1) 1992–93 NFHS: 7,815 ever-married women aged 15–19, (2) 1998–99 NFHS: 7,041 ever-married women aged 15–19, (3) 2005–06 NFHS: 23,955 women aged 15–19, (4) 2015–16 NFHS: 124,878 women aged 15–19, (5) 2019–21 NFHS: 122,480 women aged 15–19 years, and (6) Sample Registration System (SRS) from 1971 to 2007: 313 state-level ASFR 15–19 records.

**Outcome measures:** In 2025, Jammu and Kashmir had the lowest ASFR estimates at 4.0 births with a 95% credible interval [0.5; 35.7] per 1,000 females aged 15–19, while West Bengal had the highest rate at 55.4 [18.2; 157.7] births per 1,000. High 15–19 ASFR estimates concentrated in north-eastern states. We project adolescent fertility to either decline across states/UTs or remain stable in those estimated at low levels prior 2025. By 2050, the 15–19 ASFRs are projected to range from 0.7 [0.1; 3.6] to 47.7 [13.5; 150.8] per 1,000 across Indian states/UTs.

**Conclusions:** This study provides the first estimates and probabilistic projections of ASFR for ages 15–19 at the state level in India, including uncertainty estimates based on reproducible models. These outcomes can assist in region-specific policy planning to address child marriage, expand contraceptive access among adolescents, and strengthen educational and gender-equity initiatives.

**ARTICLE SUMMARY:** *Strengths and limitations of this study:*

- We developed a Bayesian spatiotemporal hierarchical modelling framework that incorporates measurement uncertainty and differences in data quality across sources, and enables information sharing across states/UTs to improve estimates in states/UTs that lack data.
- The study is based on an extensive database of state-level ASFR aged 15–19, including five India National Family Health Surveys and the Sample Registration System, totalling 1,510 fertility observations.
- By modelling key state-level covariates such as marital status (with a flexible nonlinear structure), education attainment, and total fertility, the approach captures subnational demographic variations in India.
- The modelling choice of assuming random effects for selected covariates may not fully capture state-level dynamics or shifts over time.
- The probabilistic projections are subject to model assumptions and priors that the relationships between covariates and adolescent fertility remain stable, which may be inadequate if major behavioural, policy changes, or plausible prior information occur in the future.

## INTRODUCTION

India has historical records of child marriage dating back to the early 20^th^ century.[1,2,3] The prevalence of child and early marriages in India was primarily due to the widespread practice of prepubertal betrothals and poverty.[4] Since then, the median age at marriage for women in India has gradually increased, yet it remains among the global lowest. In 2023, the median age of first marriage in India was 23, according to the Sample Registration System (SRS).[5] Women who marry early tend to give birth shortly after marriage, and about half of them undergo sterilisation before the age of 35.[6,7,8] With negligible non-marital fertility, child and early marriage are the primary drivers of adolescent fertility rates in India. According to the United Nations (UN), India surpassed China to become the world’s most populous country in 2023.[9] A good understanding and perception of the current and future levels and trends in adolescent fertility rates in India is vital in the global adolescent fertility scenario. It is also crucial for population estimates and projections in India and globally, given India’s demographic weight.

Adolescent fertility has rapidly declined worldwide and virtually disappeared in many countries. However, the number of teen births remains high in several regions, including Sub-Saharan Africa and South Asia.[10,11] Unlike other countries, India has achieved fertility decline despite persistent child marriage, early first births and the reliance on non-reversible family planning methods (viz. female sterilisation).[12] An additional challenge for understanding India’s fertility transition relates to its caste and religious diversity and the considerable demographic disparities observed across regions. Some states, like Kerala, have demographic profiles akin to those of Eastern Europe in terms of child mortality and to those of Western Europe on overall fertility according to reported values from National Family and Health Survey (NFHS; India Demographic and Health Surveys) 2015–16 and 2019–21.[6,7] Meanwhile, less developed states (with a typical demographic profile of high fertility combined with a high child mortality), like Bihar, resemble countries in Sub-Saharan Africa. Subregional mortality and fertility estimates for Bihar are reported with outlying high values compared with most parts of south India.[37] In view of its heterogeneous nature, studying India’s adolescent fertility rates necessitates a subnational approach. Although the legal marriage age for females in India is set at 18, early marriage remains prevalent while exhibiting significant variations across regions and, as indicated earlier, is the prime factor of adolescent fertility in India along with educational attainment.[13] Overlooking these regional disparities may, in turn, result in estimates or projections grossly implausible at the subnational level. Thus, a detailed, disaggregated approach to understanding the current and future development of fertility rates among females aged 15–19 is crucial for population estimation, projection, and policy planning in India.

Given the importance of a subnational perspective for estimating and projecting fertility rates in India, prior studies have addressed this issue for adolescents.[14,15,16] However, existing publications on state-level adolescent fertility rates are limited to data from a single round of the NFHS and do not analyse or model fertility trends nor do they project to the future.[17] To the best of our knowledge, this study is the first to provide estimates and projections with uncertain assessment till 2050 for the ASFR 15–19 by Indian state/union territory (UT) based on a reproducible Bayesian modelling approach.

In this paper, we estimate and project the ASFR for adolescent females aged 15–19 among 29 largest Indian states/UTs. We present state-level annual estimates and projections with uncertainty intervals from 1990 to 2050 for 21 states/UTs with population above 10 million based on the 2011 India Census, covering 91% of India’s total population as of 2023.[5] In this study, the former state of Andhra Pradesh includes Telangana (created in 2014). Similarly, Jammu and Kashmir includes Ladakh (bifurcated in 2019).

## DATA AND METHODS

The data and method details, including data preprocessing, quality assessments, model specifications, priors, statistical computations, validation exercises, and sensitivity analyses are available in the appendix (DOI:<u>10.6084/m9.figshare.30772265</u>). We provide a flowchart of data compilation and statistical modelling overview (Figure 1) and summarise the main steps in the rest of the section.

**Figure 1.**
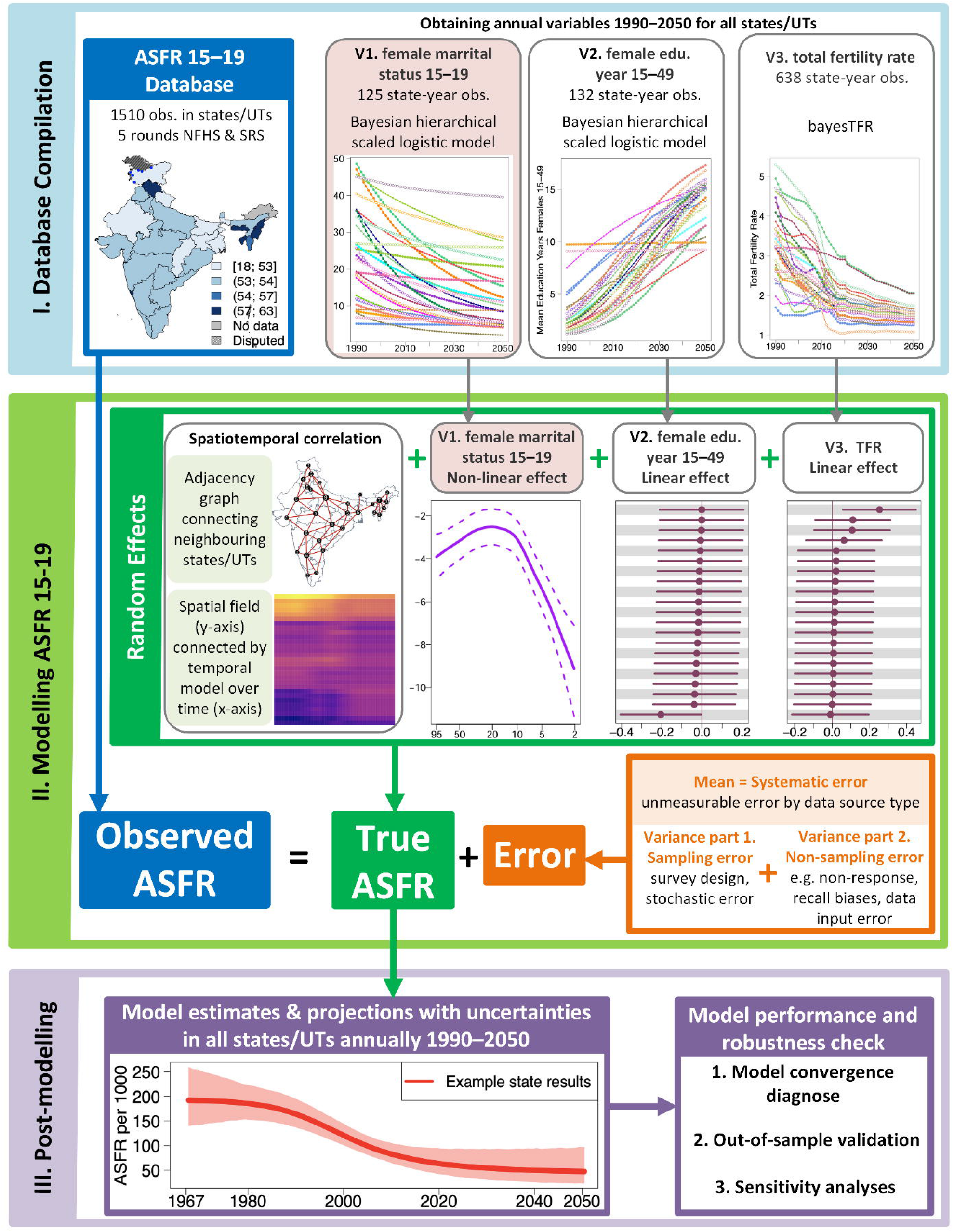
Flowchart for data and modelling process. This flowchart summarizes the major steps of data compilation (Block I), Bayesian modelling (Block II), and post-modelling (Block III). Obs.: observation(s); edu.: education. TFR: total fertility rate. V1/2/3: variable 1/2/3. Full version of all plots in the flowchart can be found in the appendix (DOI:<u>10.6084/m9.figshare.30772265</u>).

### Data

We compiled an extensive database of state-level ASFRs for 15–19-year-olds in India, comprising 1,510 observations from all states/UTs. The observation reference years range from 1967 to 2020. We processed microdata from complete birth histories collected across five rounds of NFHS, spanning 1992–1993, 1998–1999, 2005–2006, 2015–2016, and 2019–2021. We computed the corresponding sampling errors for the ASFR 15–19 observations from NFHS to reflect uncertainties arising from the multi-level stratified sample design (appendix).

The state-level covariates, serving as input variables in modelling the state-level ASFR 15–19, are specifically modeled for this study. Two covariates used in the model (mean education years among all females aged 15–49 and proportion of ever-married females aged 15–19) are estimated and projected for 1990–2050 across states/UTs using NFHS data within a Bayesian hierarchical framework (described in detail below). Total fertility rate (TFR; the third variable used in the ASFR model) data for Indian states/UTs from 1990–2050 are sourced from the India SRS and NFHS based on the bayesTFR R-package.[18] The resulting input variable estimates and projections used in the ASFR 15–19 model are provided in the appendix.

The final ASFR 15–19 database is publicly available (DOI:<u>10.6084/m9.figshare.30771611</u>) as the supplementary file. Detailed data sources and coverages for each Indian state/UT regarding the ASFR 15–19 and input data souces to obtain all covariates are available in the appendix as supplementary tables.

### Methods

We developed a Bayesian hierarchical time-series model to estimate and project the fertility rate among females aged 15–19 by Indian state/UT through 2050. The ASFR 15–19 is modelled as a combination of the effects of female marital status, educational attainment, TFR, and the spatiotemporal correlation. All the model assumptions, including the variable choice, model choice for each variable, and prior choice are validated and discussed in the appendix (pp 17–23).

#### Process model for ASFR aged 15–19

The outcome of interest is the ASFR for the age group 15–19 in Indian state/UT *s* in year *t*, modelled on the logit scale, and denoted as Ψ_*s,t*_. Let *y*_*i*_ be the *i*-th observed ASFR 15–19 on the logit scale across all state-years with data. We assume that for *i* ∈ {1, …, 1510}:

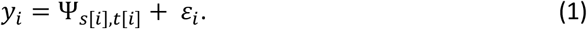

The logit-scaled observation *y*_*i*_ is modelled as the sum of: (i) Ψ_*s*[*i*],*t*[*i*]_, the true ASFR 15–19 on the logit scale in state *s*[*i*] in year *t*[*i*], and (ii) ε_*i*_, the measurement error.

We modelled Ψ_*s,t*_, the true ASFR 15–19 on a logit scale, as follows:

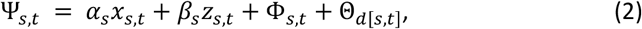

where *x*_*s,t*_ and *z*_*s,t*_ are the log of TFR and the mean education years among all females aged 15–49, respectively, for Indian state/UT *s* in year *t*. We assumed random effects for the two variables differ across Indian states/UTs, denoted as *α*_*s*_ and *β*_*s*_ for state/UT *s*. We used hierarchical normal distributions to model the two sets of state-level random effects *α*_*s*_ and *β*_*s*_.

Φ_*s,t*_ models the structured spatiotemporal fluctuations. Let Φ_*t*_ = (Φ_1,*t*_, …, Φ_29,*t*_)^𝖳^ be the spatial field vector across all states/UTs in year *t*. We used a Besag-type intrinsic conditional autoregressive (ICAR) model to capture the spatial dependency among neighbouring states/UTs in year *t*, where neighbouring states/UTs are defined as sharing states/UT boarders.[38] Using an adjacency graph which identify neighbouring states/UTs, the Besag model pools the effects from adjacent states/UTs towards each other. Across year *t*, the correlation between the year-specific spatial fields Φ_*t*_ is modelled with a first-order autoregressive (AR1) time series process such that

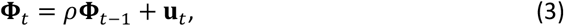

where *ρ* is the autoregressive correlation parameter, and **u**_*t*_ a spatially structured innovation term that follows the adjacency graph. The temporal component allows the spatial pattern to evolve smoothly over time while preserving the spatial autocorrelation among neighbouring states/UTs.

Θ_*d*[*s,t*]_ captures the overall non-linear relationship between Ψ_*s,t*_ and the proportion of ever-married females aged 15–19 in Indian state/UT *s* in year *t*, denoted as 𝑣_*s,t*_. Index *d*[*s, t*] denotes matching 𝑣_*s,t*_ to the *d*-th knot where Θ is evaluated. We used a second-order random walk (RW2) process to model Θ_*d*_:

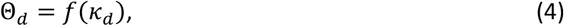

where *κ*_*d*_ for *d* ∈ 1, …, 1000 are the 1,000 knots where we modelled Θ. We matched each 𝑣_*s,t*_ to its closest *κ*_*d*_ and fitted the overall RW2 function *f*(⋅). The RW2 model is flexible and allows for potential non-linear effects from this variable, as it shows the strongest association among all three variables with the ASFR 15–19, based on exploratory analysis (see appendix p5 for illustration of the motivation for variable modelling choice).

#### Data model for ASFR aged 15–19

The error term ε_*i*_ follows a normal distribution with a mean at λ_*p*[*i*]_, the data source-specific random effect for *p* ∈ {NFHS, SRS}. The source-specific random effect λ_*p*_ models the underlying systematic discrepancies in NFHS and SRS that are unmeasurable in the data pre-processing. Its variance is assumed to be the sum of: (i) known stochastic/sampling error variance 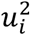 for the *i*-th observation, and (ii) unknown non-sampling error variance 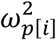 for the *p*-th data type to which the *i*-th observation belongs. Particularly,

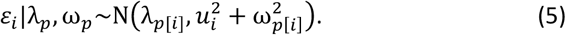

The sampling variance 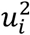 is pre-calculated using the Jackknife resampling method (detailed in appendix) [19] to account for the uncertainty due to the survey sample design or the underlying stochastic errors of the birth occurrences. The non-sampling error variances 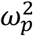 are assumed to differ across data sources NFHS and SRS. 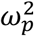 for *p* ∈ {NFHS, SRS} reflects the uncertainty due to non-response, data-entry errors, recall biases, and so forth, which can be minimised but are challenging to be eliminated. Given this data model, the Bayesian estimates are pooled towards informative observations and are less influenced by weakly informative observations.

#### Model for variables as ASFR model inputs

We obtained *z*_*s,t*_ (mean education years among all females aged 15–49) and 𝑣_*s,t*_ (proportion of ever-married females aged 15–19) for all Indian state-years from 1990 to 2050 based on scaled Bayesian hierarchical logistic models with the general structure (see appendix for model details on obtaining *z*_*s,t*_ and 𝑣_*s,t*_):

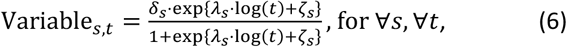

where Variable_*s,t*_ is the model fittings for the two variables *z*_*s,t*_ and 𝑣_*s,t*_. Variable_*s,t*_ is modelled with an independent variable the log of time index log(*t*), and state-specific parameters that define the features of a logit curve, including coefficients λ_*s*_ (as the rate of decline), intercept ζ_*s*_ (as the baseline level), and the scale δ_*s*_ (the maxima) for each state *s*. All state-specific parameters were assigned hierarchical distributions.

#### Post-model processing

The trajectories of the model variables were used to produce the final uncertainty intervals of the estimated ASFRs for all state-years (appendix pp 13).

#### Model validation

We assessed model performance using out-of-sample validation exercises (appendix pp 13–14). Due to the retrospective nature of the fertility data and the presence of time-series data, we left out 261 observations collected since 2016 as the testing dataset, representing 17% of the total observations, with the remaining 83% datapoints as the training dataset. This validation approach has been used in previous global health and population studies.[20,21,22,23] The validation and simulation results suggested that our model was reasonably well-calibrated with conservative predition intervals.

#### Statistical computation

All data processing, analysis, and Bayesian modelling were performed using the open-source software R 4.4.0.[24] We used the Integrated Nested Laplace Approximation (INLA) for Bayesian inference of the state-level ASFR.[25] We use the R-package R-INLA to implement the INLA estimation procedure.[26] For models of the mean education years among all females aged 15–49 and the proportion of married females aged 15–19 for all Indian state-years, we obtained posterior samples of all model parameters and hyperparameters using a Markov chain Monte Carlo (MCMC) algorithm, implemented in the open-source software JAGS 4.3.2 (Just another Gibbs Sampler),[27] with R-packages R2jags[28] and rjags.[28,29] The convergence of the MCMC algorithm and the sufficiency of the number of samples obtained were checked through visual inspection of trace plots and convergence diagnostics of Gelman and Rubin,[30] implemented in R-package coda.[31]

### Patient and Public Involvement

Patients or the public were not involved in the design, or conduct, or reporting, or dissemination plans of our research.

### Research Ethics Approval: Human Participants

This study does not involve human participants.

## RESULTS

The ASFR 15–19 annual estimates and projections from 1990 to 2050, including uncertainty for the 21 Indian states/UTs, are presented in the appendix as supplementary figures. We focus on results in 1990, 2025, and 2050 here.

### ASFR estimates in 2025 by Indian state/UT

The levels and trends in the ASFR 15–19 estimates from 1990 to 2025 vary across Indian states/UTs (Table 1). In 2025, the ASFR ranges from 4.0 (95% credible interval [0.5; 35.7]) per 1,000 females aged 15–19 in Jammu and Kashmir to 55.4 [18.2; 157.7] per 1,000 in West Bengal.

**Table 1.** Estimates and 95% uncertainty intervals for ASFR 15–19 in 1990, 2025, and 2050, by Indian state/UT. †: total reduction or annual rate of reduction (ARR) is significantly different from zero. Total reduction: difference between values at start and end of the period. Annual rate of reduction: ln 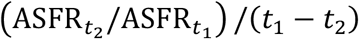, where *t*_1_ and *t*_2_ refer to different years with *t*_1_ smaller than *t*_2_, and 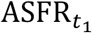 and 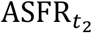 are ASFR 15–19 in years *t*_1_ and *t*_2_ respectively.

| Age-specific Fertility Rate 15–19 |  |  |  |  |  |  |  |
| --- | --- | --- | --- | --- | --- | --- | --- |
| Indian state/UT Name | Estimate and Projection (per 1,000 females) |  |  | Total Reduction (per 1,000 females) |  | Annual Rate of Reduction (in %) |  |
|  | 1990 | 2025 | 2050 | 1990–2025 | 2025–2050 | 1990–2025 | 2025–2050 |
| former state of Andhra Pradesh | 119.2<br>[36.4; 338.6] | 34.3<br>[8.7; 127.1] | 15.3<br>[3.0; 75.6] | 84.9<br>[-22.3; 289.7] | 19.1<br>[-25.6; 93.3] | 3.56<br>[-0.92; 8.06] | 3.25<br>[-2.98; 9.67] |
| Assam | 91.3<br>[33.0; 222.7] | 31.2<br>[10.1; 94.6] | 20.6<br>[5.9; 69.5] | 60.1<br>[-18.4; 188.6] | 10.6<br>[-34.4; 65.2] | 3.07<br>[-0.92; 6.94] | 1.65<br>[-3.91; 7.30] |
| Bihar | 102.3<br>[30.4; 287.6] | 25.6<br>[7.4; 85.2] | 8.2<br>[2.0; 32.6] | 76.7<br>[-4.1; 251.3] | 17.4<br>[-4.9; 70.7] | 3.96<br>[-0.22; 7.95] | 4.54<br>[-1.18; 10.57] |
| Chhattisgarh | 105.5<br>[36.4; 264.7] | 12.2<br>[3.9; 38.5] | 5.4<br>[1.5; 19.5] | 93.3<br>[22.8; 249.2]† | 6.9<br>[-5.7; 30.0] | 6.15<br>[2.16; 9.96]† | 3.30<br>[-2.14; 8.90] |
| Delhi | 49.8<br>[6.9; 267.1] | 12.2<br>[2.2; 64.1] | 6.5<br>[0.9; 45.0] | 37.6<br>[-15.5; 237.3] | 5.6<br>[-20.6; 45.0] | 4.03<br>[-1.85; 9.46] | 2.48<br>[-4.71; 9.67] |
| Gujarat | 68.5<br>[21.9; 196.4] | 21.1<br>[6.2; 67.3] | 11.4<br>[3.0; 43.5] | 47.4<br>[-10.6; 168.8] | 9.7<br>[-18.1; 50.0] | 3.37<br>[-0.78; 7.45] | 2.46<br>[-3.39; 8.30] |
| Haryana | 79.9<br>[26.5; 220.2] | 9.2<br>[2.6; 32.4] | 0.7<br>[0.1; 3.6] | 70.6<br>[15.8; 205.4]† | 8.5<br>[2.0; 31.1]† | 6.16<br>[1.92; 10.39]† | 10.30<br>[3.95; 16.64]† |
| Himachal Pradesh | 57.6<br>[18.1; 173.3] | 7.4<br>[1.8; 29.9] | 2.7<br>[0.5; 15.1] | 50.2<br>[7.7; 162.2]† | 4.7<br>[-4.9; 23.7] | 5.85<br>[1.14; 10.53]† | 4.05<br>[-2.86; 10.75] |
| Jammu and Kashmir | 55.0<br>[7.0; 304.1] | 4.0<br>[0.5; 35.7] | 1.4<br>[0.1; 25.1] | 51.0<br>[-0.3; 292.0] | 2.6<br>[-13.9; 28.2] | 7.49<br>[-0.05; 14.19] | 4.06<br>[-7.02; 15.34] |
| Jharkhand | 92.9<br>[31.7; 251.0] | 34.0<br>[10.6; 106.3] | 19.8<br>[5.8; 64.9] | 58.9<br>[-18.8; 202.8] | 14.2<br>[-24.9; 74.0] | 2.87<br>[-0.95; 6.64] | 2.17<br>[-3.14; 7.55] |
| Karnataka | 105.2<br>[31.1; 293.6] | 20.2<br>[5.3; 73.4] | 5.1<br>[1.0; 26.7] | 85.0<br>[4.5; 264.1]† | 15.1<br>[-4.9; 62.7] | 4.71<br>[0.31; 9.14]† | 5.51<br>[-1.44; 12.37] |
| Kerala | 43.3<br>[10.4; 164.3] | 4.5<br>[0.8; 25.4] | 0.9<br>[0.1; 6.7] | 38.9<br>[5.2; 154.6]† | 3.6<br>[-0.7; 22.5] | 6.49<br>[1.23; 11.61]† | 6.57<br>[-1.18; 14.18] |
| Madhya Pradesh | 112.9<br>[37.0; 287.2] | 18.0<br>[5.6; 56.3] | 9.3<br>[2.5; 33.6] | 94.9<br>[15.2; 263.4]† | 8.6<br>[-12.2; 41.1] | 5.25<br>[1.09; 9.17]† | 2.63<br>[-3.06; 8.05] |
| Maharashtra | 98.1<br>[32.4; 263.3] | 24.1<br>[7.4; 74.5] | 10.8<br>[2.5; 42.5] | 74.0<br>[-0.9; 228.8] | 13.3<br>[-14.5; 58.2] | 4.01<br>[-0.07; 7.96] | 3.20<br>[-2.69; 9.26] |
| Odisha | 83.3<br>[28.8; 216.2] | 15.6<br>[4.9; 48.7] | 6.8<br>[1.7; 25.5] | 67.7<br>[7.6; 196.2]† | 8.8<br>[-7.4; 37.5] | 4.79<br>[0.69; 8.76]† | 3.30<br>[-2.29; 8.87] |
| Punjab | 49.7<br>[15.9; 147.5] | 8.8<br>[2.1; 35.7] | 3.2<br>[0.5; 17.9] | 40.9<br>[2.0; 134.7]† | 5.6<br>[-5.4; 27.8] | 4.96<br>[0.27; 9.66]† | 4.08<br>[-2.54; 10.98] |
| Rajasthan | 92.6<br>[29.7; 253.3] | 16.1<br>[5.1; 52.6] | 8.5<br>[2.3; 30.8] | 76.4<br>[9.6; 229.4]† | 7.6<br>[-10.7; 37.8] | 5.00<br>[0.82; 9.00]† | 2.56<br>[-2.90; 7.93] |
| Tamil Nadu | 71.4<br>[21.1; 215.8] | 10.2<br>[2.2; 46.2] | 1.3<br>[0.2; 8.5] | 61.2<br>[7.0; 196.9]† | 8.9<br>[0.8; 42.6]† | 5.55<br>[0.77; 10.23]† | 8.16<br>[1.14; 14.87]† |
| Uttar Pradesh | 94.1<br>[33.7; 236.5] | 11.8<br>[4.0; 33.5] | 5.5<br>[1.7; 17.1] | 82.3<br>[20.2; 222.7]† | 6.3<br>[-5.5; 25.6] | 5.93<br>[2.18; 9.64]† | 3.05<br>[-2.30; 8.25] |
| Uttarakhand | 63.5<br>[7.9; 398.8] | 15.0<br>[2.3; 92.1] | 8.2<br>[1.0; 60.9] | 48.5<br>[-12.5; 356.4] | 6.8<br>[-26.0; 67.6] | 4.12<br>[-1.39; 9.71] | 2.41<br>[-4.92; 10.19] |
| West Bengal | 103.9<br>[37.6; 250.6] | 55.4<br>[18.2; 157.7] | 47.7<br>[13.5; 150.8] | 48.5<br>[-61.4; 191.0] | 7.8<br>[-81.7; 94.2] | 1.79<br>[-2.11; 5.52] | 0.60<br>[-4.55; 5.90] |

During the period 1990–2025, all 21 Indian states/UTs are estimated to experience decreases in the ASFR 15–19. 11 states/UTs experienced total reductions from 1990 to 2025 that are significantly different from zero. The largest absolute reduction is estimated in Madhya Pradesh at 94.9 [15.2; 263.4] per 1,000 and Chhattisgarh at 93.3 [22.8; 249.2] per 1,000, and the smallest decline is in Delhi at 37.6 [-15.5; 237.3] per 1,000. On a relative scale, the decline in adolescent birth rates is measured by the annual reduction rate (ARR), where a positive ARR indicates a decrease in the teenage birth rate during the period. Jammu and Kashmir achieved the fastest annual rate of reduction, with its ARR estimated at 7.49% [-0.05%; 14.19%] from 1990 to 2025, alongside 6.16% [1.92%; 10.39%] in Haryana and 6.15% [2.16%; 9.96%] in Chhattisgarh.

Geographically, we estimate that ASFR 15–19 varies greatly across Indian states/UTs in 2025 (Figure 2). The highest state-level adolescent fertility rates are concentrated in most of the north-eastern states/UTs, such as Bihar, Jharkhand, and West Bengal. The high-ASFR belt extends slightly towards the south, except for Kerala at the southern end of the Indian peninsula. The estimated ASFR 15–19 decreases for states/UTs further north.

**Figure 2.**
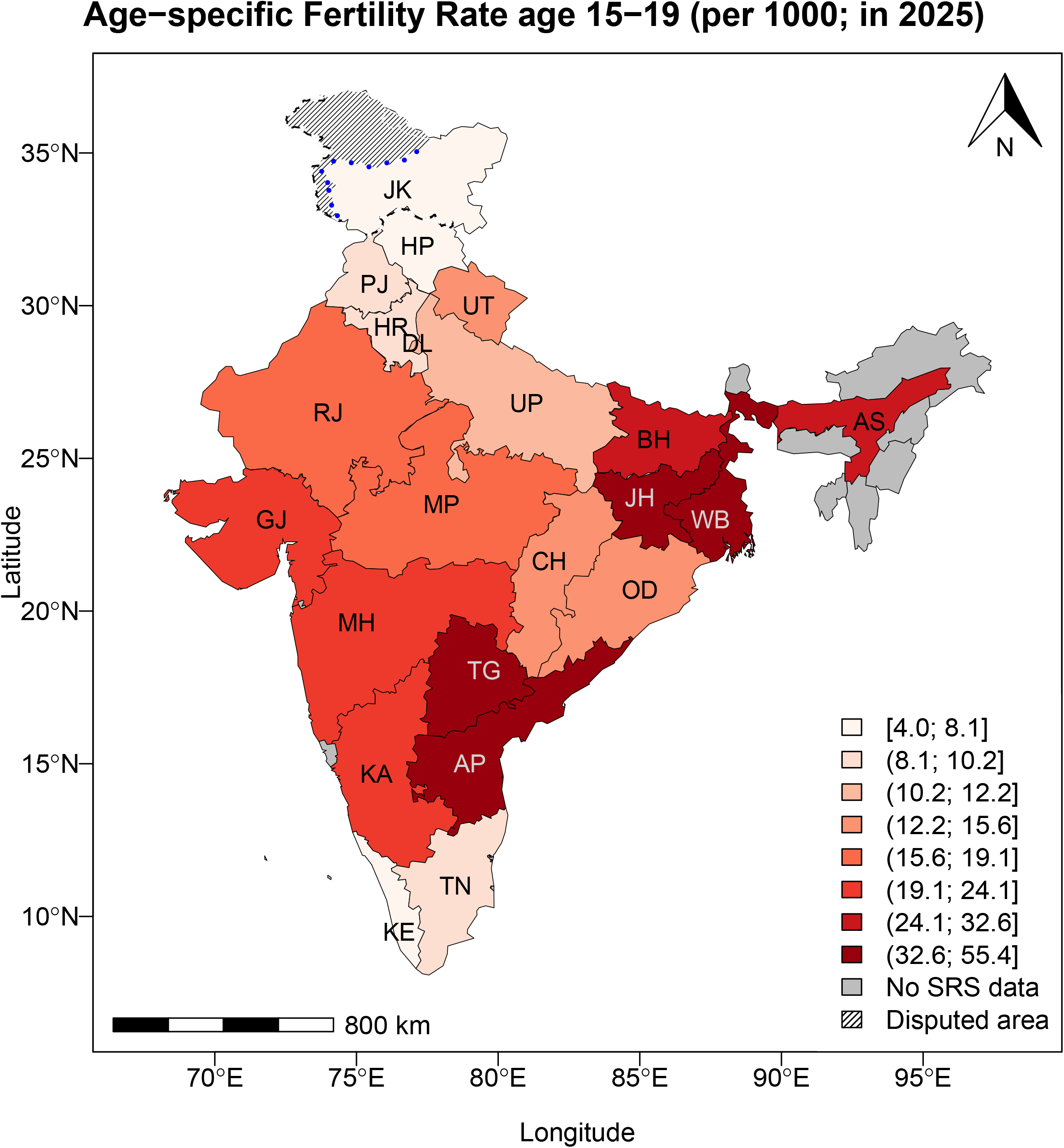
ASFR 15–19 median estimates in 2025, by Indian state/UT. Results are shown for the 21 Indian States/UTs with SRS data. Values for Andhra Pradesh and Telangana are the same. State/UT names are: Andhra Pradesh (AP); Assam (AS); Bihar (BH); Chhattisgarh (CH); Delhi (DL); Gujarat (GJ); Haryana (HR); Himachal Pradesh (HP); Jammu and Kashmir (JK); Jharkhand (JH); Karnataka (KA); Kerala (KE); Madhya Pradesh (MP); Maharashtra (MH); Odisha (OD); Punjab (PJ); Rajasthan (RJ); Tamil Nadu (TN); Telangana (TG; same value as AP); Uttar Pradesh (UP); Uttarakhand (UT); West Bengal (WB). The boundaries and names shown, as well as the designations used on this map, do not imply official endorsement. The blue dotted line represents approximately the Line of Control in Jammu and Kashmir, as agreed upon by India and Pakistan. The parties have not yet agreed upon the final status of Jammu and Kashmir. The scale bar may have minor inaccuracies.

### ASFR projections for 2025–2050 by Indian state/UT

Figure 3 shows the ASFR 15–19 model estimates and projections by Indian state from 1990 to 2050 among the 21 largest Indian states/UTs. The projection period is zoomed in on the bottom panel of Figure 3. By 2050, projections range from 0.7 [0.1; 3.6] per 1,000 in Haryana to 47.7 [13.5; 150.8] per 1,000 in West Bengal (Table 1).

**Figure 3.**
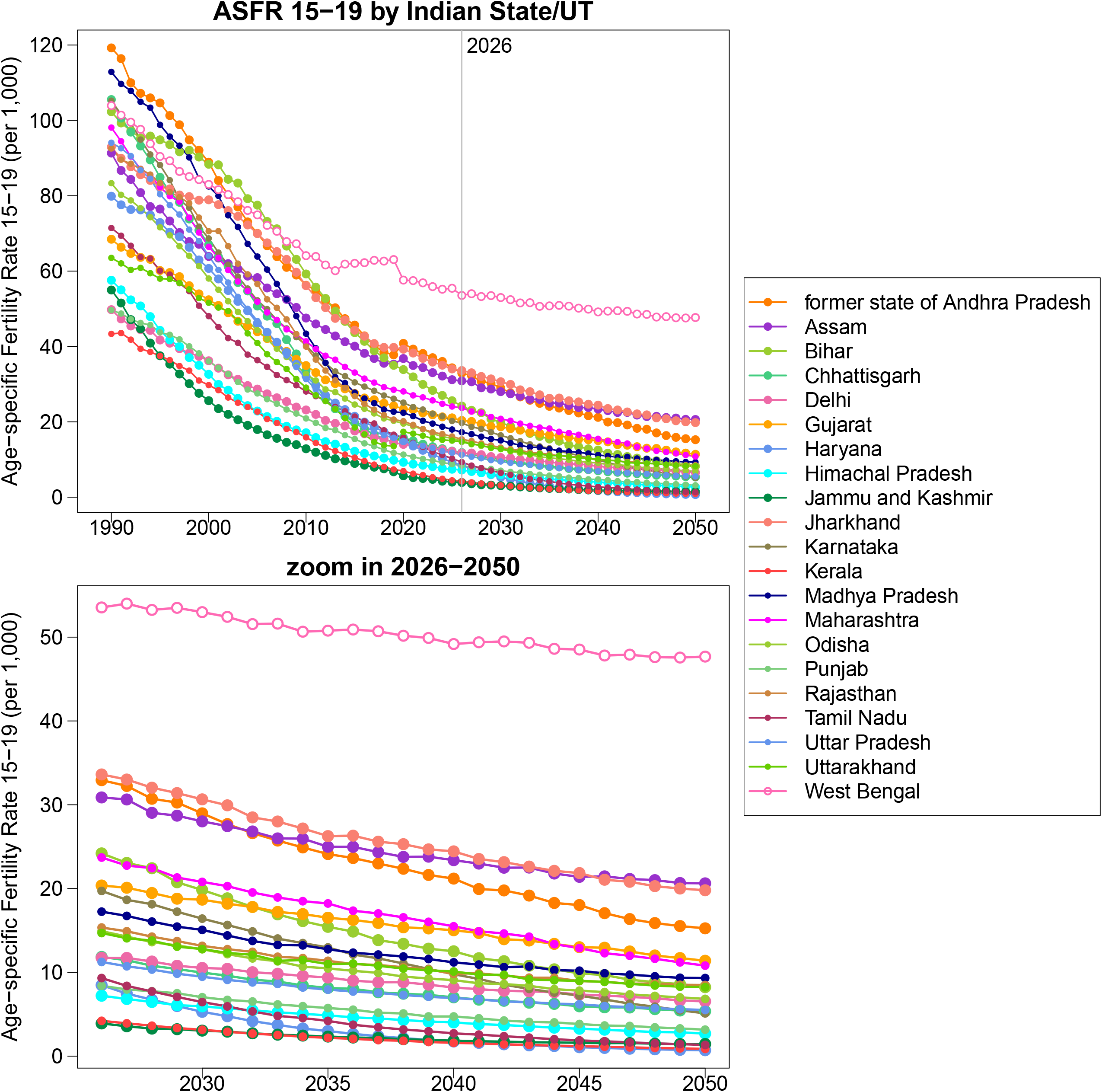
ASFR 15–19 model estimates and projection by Indian state/UT from 1990 to 2050. Median estimates and projections are shown. States/UTs are differentiated by color. Results are presented for the 21 largest Indian states/UTs.

The Bayesian model results imply a steady decline in ASFR 15–19 across states/UTs, even at current low levels in 2025. For states/UTs with the highest teenage fertility currently, such as West Bengal, Assam, and Jharkhand, their 2050 ASFR 15–19 are projected to remain the largest among Indian states/UTs, with moderate ARRs projected in these states/UTs (Table 1). Most states with low teenage ASFRs in 2025 are projected to maintain their current levels till 2050. For example, Haryana, Kerala, Jammu and Kashmir, Himachal Pradesh, and Tamil Nadu have the lowest ASFRs in both 2025 and 2050. All three low-fertility states/UTs are projected to experience the fastest decline during the projection period 2025 to 2050.

### State-specific case study

The ASFR 15–19 estimates and projections for three selected Indian states/UTs, as shown in Figure 4, illustrate the diversity of ASFR 15–19 trajectories in India. Figure 4 presented the period prior 1990 as long as is covered with data.

**Figure 4.**
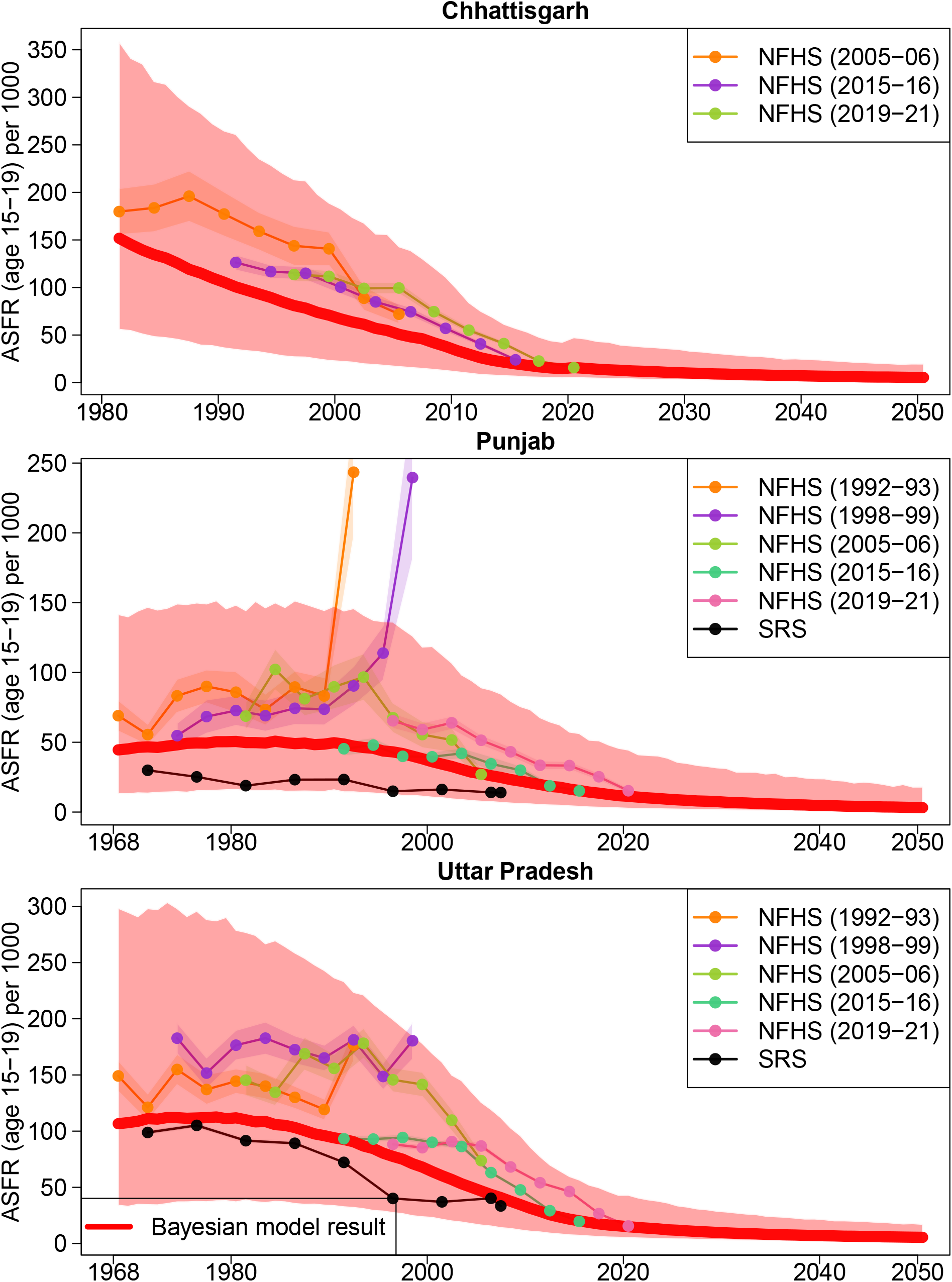
ASFR 15–19 in selected Indian states/UTs. Red curves: posterior medians. Red shades: 95% credible intervals. Dots with connection lines: input data series differentiated by colors. Shades around the data series: sampling/stochastic errors.

The first case is Chhattisgarh, which has one of the highest absolute declines in ASFR, with its 2025 level 93.3 [22.8; 249.2] per 1,000 lower than in 1990. We estimate that the ASFR 15–19 in Chhattisgarh declined monotonically from 157.7 [57.2; 361.2] per 1,000 in 1980 to 12.2 [3.9; 38.5] per 1,000 by 2025. Our model projections suggest that the fertility rate among adolescent females aged 15–19 will have mild decline and reach at 5.4 [1.5; 19.5] per 1,000 in 2050. This moderate projected decline is because the proportion of married teenage females in this region has reached a low level before 2025, and projections are almost constant for most periods.

Punjab has had historically low fertility rates since the 1968, the earliest year in which data are available. Punjab is unique, as it shows a slight “humped” pattern in ASFR 15–19 in the 1970s and 1980s. The ASFR 15–19 in Punjab is estimated to have continued to grow from 44.5 [13.2; 141.5] per 1,000 in 1968 to 49.7 [15.9; 147.5] per 1,000 in 1990, although the increase is not statistically significant. Same as in Chhattisgarh, the ASFR 15–19 projections in Punjab remain moderate and stable, primarily driven by the proportion of married females in this age group.

Uttar Pradesh is of primary importance, as it is the most populous state in India, with an estimated population of 237 million as of 2020. The ASFR 15–19 in Uttar Pradesh follows a typical pattern in states/UTs with good data coverage dated back to the 1960s where the ASFR 15–19 remain stable and starts to decline at a faster speed over time; its ASFR 15–19 was at 106.2 [34.0; 298.3] per 1,000 in 1968 and start to decline noticeably in mid 1980s. The ASFR 15–19 is estimated to decrease to 11.8 [4.0; 33.5] per 1,000 till 2025. The model projects a slower decline in ASFR 15–19 in Uttar Pradesh to 5.5 [1.7; 17.1] per 1,000 by 2050. The former state of Andhra Pradesh and Madhya Pradesh show similar downward trends, which is crucial for national adolescent fertility rates since these states contribute to roughly 15% births in the country in 2025.[32]

## DISCUSSION

This study is the first to provide state-level estimates and probabilistic projections of adolescent fertility rates for ages 15–19 in India till 2050. Our ASFR 15–19 model results are essential for population estimation and projection models in India, particularly at the sub-national level. We incorporated measurement uncertainty through a Bayesian modelling framework, ensuring that varying data qualities are reflected in the model output in a reproducible fashion. Furthermore, the hierarchical model structure captures regional diversity and spatiotemporal correlation in adolescent fertility patterns and preserves subnational demographic disparities in India.

Our Bayesian modelling approach accounts for the teenage female marital status, female educational attainment, and TFR, each playing a vital role in shaping adolescent fertility levels and trends. The choice of covariates in our model depends on their ability to approximate the effects of fertility decisions among females together with the reliability and availability of their estimates. We allow for potential non-linear effects in the proportion of married females aged 15–19, as it shows the strongest correlation (linear or non-linear) in our exploratory analysis (appendix). Assuming potential nonlinearity in the proportion of married adolescent Indian females enables the model to better capture the underlying dynamics between teenage female marital status and corresponding fertility patterns. The early initiation of sexual activity, coupled with societal pressure on young married women to demonstrate fertility shortly after marriage in India, is reflected in certain states/UTs where early marriage triggers elevated adolescent fertility rates. Demographic surveys such as the NFHS (phases II to V) reveal alarmingly low contraceptive use among teenage women, resulting in many young women becoming mothers shortly after marriage.[1,2,6,7,8]

Our study has limitations arising from data processing and Bayesian model assumptions, which are inherent to all modelling studies. First, it is important to note that the estimation results prior to 2025 in state-periods without observations are based on model assumptions derived from the universal set of predictors used in the Bayesian model. While the current variable choices may work well in most Indian states/UTs, it may not apply to all. Second, we focus on modelling the effect of marital status on adolescent fertility, assuming that the effects of educational attainment and total fertility are linear based on empirical evidence from NFHS data. This may not fully reflect the reality across all Indian states/UTs, as education and/or total fertility might play a stronger role in some regions. Third, readers should be cautious when interpreting probabilistic projection results, as the underlying projection assumption is that all other conditions remain constant while controlling for the effects included in the Bayesian model. Furthermore, although the ASFR model is robust during the estimation period prior 2025 against the prior choices, the projections are subject to a certain level of sensitivity; under certain prior scenarios (either more vague or more informative than the default model setup), the projections and associated uncertainty bounds are marginally affected (appendix pp 20–23). Fourth, the consistent discrepancy between the observations from the SRS and NFHS may imply potential birth underreporting in SRS and/or backward displacement of births in the NFHS 1992–93.[36] However, these speculations are not further discussed anywhere else nor is there study about more recent fertility data comparison. Given the lack of evidence to discard or adjust data by source type, we modelled the data source random effects instead. Fifth, combining the two-step modelling into one model with a fully Bayesian approach is not feasible, because the model became computational costly and unable to achieve convergence. Consequently, the modelling process has to be carried out in two steps to produce reasonable results. All the aforementioned limitations can be addressed through in-depth studies if plausible information on the predictors or high-quality observations of ASFR 15–19 and associated covariates become available in the future for some Indian states/UTs.

India is unique in its regional diversity of ASFR trajectories among 15–19-year-olds. In states like West Bengal, the former state of Andhra Pradesh, Bihar, and Jharkhand, which had historically high levels of adolescent fertility in 1990 and are projected to maintain the highest levels in 2025 with moderate declines (measured by the ARR), early marriage remains prevalent.[5,33] In these Indian states/UTs, adolescent fertility accounts for over 15% of total fertility rates. Given the diversity of regional circumstances, a one-size-fits-all approach may be ineffective, and hence our state-level method filled the need of addressing the heterogeniety.

In states like Bihar and Jharkhand, high adolescent birth rates are part of a patriarchal reproductive system where early marriage and low contraceptive rates contribute to some of the highest TFR levels in India. Poverty and low female educational attainment are central to this traditional fertility system. In these states, policy priorities should continue to address all aspects of reproductive health and gender equity. The specific challenges faced by young women married during adolescence should be addressed through initiatives that empower girls and expand secondary education and vocational training to enhance their potential as income earners. Campaigns should also target the traditional norms favouring child marriage. These transformations will ultimately change the cost-benefit ratio of early marriage.

In contrast, the adolescent fertility trend in West Bengal and the former state of Andhra Pradesh remains directly linked to the persistence of early marriage among a significant portion of the population–despite recent advances in fertility reduction and women’s education. The fertility window for married women in these regions is compressed, resulting in an unhealthy combination of early marriage, childbearing, and sterilisation.[4] Policy focus in these regions should be on preventing child marriage through normative changes and expanding access to reversible contraceptives to delay the first birth and extend the reproductive span beyond the mid-twenties. Public health communication should target adolescents and young adults, shifting from discussions about the number of children (which is already below replacement level) to delaying marriage and timing the first pregnancy.

More affluent states/UTs, such as Kerala, Himachal Pradesh, Delhi, and Punjab, are now experiencing fewer teenage births. For example, child marriage has become uncommon in Punjab and the contraception rate has reached three-quarters of married women aged 15–19 since 2015.[6] Access to education and to pre-conception counselling tailored for young married women plays a pivotal role in these regions, serving as both a cause and a consequence of high contraception coverage among married teenage females.[34]

Adolescent fertility poses potential burdens and risks for both individuals and the population. First and foremost, adolescent females face substantial challenges related to their sexual and reproductive health and rights. Pregnancy during adolescence often occurs before individuals are fully developed, exposing them to significant health risks during pregnancy and childbirth. The nutritional demands of pregnancy coincide with the adolescent growth spurt, a phase that already necessitates increased nutritional intake. Inadequate nourishment can exacerbate challenges faced by malnourished adolescents. Early pregnancy can severely damage the reproductive tract, increase maternal mortality risks, lead to complications during pregnancy, and increase perinatal and neonatal mortality rates, as well as low birth weight. Adolescents are more likely to experience adverse pregnancy outcomes, such as stillbirths, miscarriages, anaemia, and weight loss, in comparison to older women. Children born to adolescent mothers are also likely to suffer from higher death rates in infancy and childhood, and to exhibit higher rates of stunting. Secondly, on the socioeconomic and public health prospects, teenage mothers are at a greater risk of experiencing social and economic disadvantages throughout their lives compared to those who delay childbearing until their twenties, potentially leading to generational poverty. They are less likely to complete their education, face reduced employment opportunities, earn lower wages, and tend to have larger families. Additionally, they are more susceptible to unprotected pregnancies and sexually transmitted infections, often lacking decision-making power in their sexual relationships.[35]

Our Bayesian probabilistic estimates and projections of ASFR 15–19 by Indian state/UT underscore the importance of monitoring and projecting the ASFR 15–19 over time at the subnational level, especially in India, the most populous country globally within a highly heterogeneous demographic context. Thus, even with limited healthcare resources, we can better monitor and safeguard the rights of adolescent girls through improved identification, monitoring, and education in the worst-affected regions. Our study highlights the need to strengthen policies that promote gender equity in access to more life choices and to introduce support measures to counter existing gender biases, tailored to each regional context. Future work may include additional sources of heterogeneity, such as education, religion, and ethnicity, to estimate ASFR 15–19 in India and ultimately to predict it.

## Data Availability

All data produced in the present work are contained in the manuscript and are available online at figshare.

https://doi.org/10.6084/m9.figshare.30771611

https://doi.org/10.6084/m9.figshare.30772265

## ETHICS STATEMENTS

### Patient consent for publication

Not applicable.

## ACKNOWLEDGEMENTS

We thank Qiqi Qiang and Yichang Shi for their valuable comments and discussion on the earlier version of this manuscript. We also thank Hana Sevcikova for the helpful guidance on using the R-package bayesTFR. We acknowledge the copy-editing by Weichu Zhao and flowchart improvement from Yiwen Li. We appreciate the insightful comments and in-depth discussions from the reviewers on the earlier versions of this manuscript. We are grateful to the DHS Program for making the very informative NFHS data freely available to registered users.

## Author contributions

FC proposed and conceptualised the study, oversaw the study design, developed the statistical model, and wrote the first draft of the manuscript and the technical appendix. FC, YS, and CZG constructed the covariate and ASFR databases. FC and YS revised the technical appendix. CZG provided policy implications. FC and CZG analysed the results and revised the manuscript. All authors edited the manuscript, read, and agreed on the final manuscript.

## Funding

FC is supported by the start-up research grant UDF01003604 from The Chinese University of Hong Kong, Shenzhen. The funder did not influence the results/outcomes of the study despite the author’s affiliation with the funder.

## Competing interests

None.

## Data sharing statement

The technical appendix is available from the figshare repository, DOI:<u>10.6084/m9.figshare.30772265</u>. The database for ASFR 15–19 by Indian state/UT is available from the figshare repository, DOI:<u>10.6084/m9.figshare.30771611</u>. The NFHS microdata files are available from the DHS Program website.

## Patient consent for publication

Not required.

## Notes

### Competing Interest Statement

The authors have declared no competing interest.

### Funding Statement

This study was funded by The Chinese University of Hong Kong, Shenzhen.

### Author Declarations

The study used (or will use) ONLY openly available aggregated human data on the population level that were originally located at DHS Program and India SRS reports.

### Summary of Updates

Model updated by adding in the spatial correlation. All results, including the table and figures are updated accordingly due to the update in the model. More discussions are added, especially on the study limitations.

